# Automated MRI-Based Segmentation of the Cranio-Cervical Venous System Using a Dedicated Deep Learning Pipeline

**DOI:** 10.64898/2026.11.08.26367527

**Authors:** Marie Duigou, Timothé Treguer, Brieg Dissaux, Julien Ognard, Romuald Seizeur, Cyrille Capel

## Abstract

**PURPOSE:** To develop an automated magnetic resonance imaging based segmentation pipeline for the cranio-cervical venous system (CCVS), a complex anatomical network for which no dedicated segmentation tool is currently available.

**MATERIALS AND METHODS:** A retrospective single-center cohort of 194 contrast-enhanced cranio-cervical MRI examinations acquired between 2019 and 2024 was used. Manual reference segmentations were initially performed on 50 examinations using 3D Slicer. A nnU-Net v2-based segmentation model was then trained through an iterative process combining automatic segmentation, manual correction, and retraining. A dedicated intermediate model was developed to distinguish the vertebral artery from the surrounding vertebral venous plexus. Segmentation performance was assessed using the Dice Similarity Coefficient (DSC). The complete processing pipeline was integrated into an open-source software application, VenousNetworkAnalyzer (VNA).

**RESULTS:** The final model achieved a mean DSC of 0.94 (95% CI, 0.93–0.95) in five-fold cross-validation on 106 examinations with manual reference segmentations. Iterative optimization progressively reduced manual processing time from approximately 10 hours of fully manual segmentation to less than 10 minutes of manual correction per patient. Application of the final model to the entire cohort enabled the generation of patient-specific three-dimensional reconstructions of the cranio-cervical venous system. The segmentation workflow was successfully integrated into the VNA software environment, allowing automated processing and visualization of MRI datasets.

**CONCLUSION:** This study demonstrates the feasibility of automated segmentation of the cranio-cervical venous system from contrast-enhanced MRI using a dedicated deep-learning pipeline. The resulting reduction in processing time and integration into an open-source software platform provide a foundation for future large-scale anatomical studies of cranio-cervical venous drainage.

**Summary Statement:** An automated nnU-Net-based MRI pipeline enabled segmentation and three-dimensional visualization of the cranio-cervical venous system, reducing manual correction time less than 10 minutes per case.

**Key Points:**

- A dedicated nnU-Net v2-based pipeline was developed for MRI-based segmentation of the cranio-cervical venous system
- Iterative model optimization reduced manual processing time from approximately 10 hours to less than 10 minutes per case.
- The pipeline enabled patient-specific three-dimensional reconstructions and was integrated into a dedicated software environment for visualization and analyzis.

## INTRODUCTION

The craniocervical junction is a major anatomical crossroads housing a complex venous system: the craniocervical junction venous system (CCVS)^1,2^. This system includes the vertebral artery venous plexus, epidural venous plexuses, perimedullary veins, deep cervical veins, and muscular and subcutaneous venous networks^1,2^.

Previous anatomical studies have mainly focused on isolated components of the CCVS. However, no comprehensive characterization of this system based on imaging is currently available. The segmentation of the CCVS on medical imaging presents a significant technical challenge. The structures to be analysed are numerous, tortuous, of varying caliber, and located in an anatomical region where arterial, venous, bony and muscular structures coexist in close proximity^1–5^. Manual segmentation allows for precise analysis, but it is time-consuming and difficult to apply to large cohorts. Although vascular segmentation in medical imaging has seen significant advances, currently available methods remain poorly suited to the CCVS^6^. These limitations prompted the development of an automatic segmentation method specifically designed for the CCVS.

Beyond this technical challenge, a more comprehensive characterization of the CCVS is also anatomically and clinically relevant. The classical dichotomous model of cerebral venous drainage between the internal jugular and vertebral venous pathways may be overly simplistic, as suggested by reports of bilateral internal jugular vein ligation without major neurological consequences. These observations support the existence of additional, functionally relevant cervical venous pathways that remain incompletely characterized.

The main objective of this study was to develop an automatic segmentation model of the CCVS based on contrast-enhanced 3D T1 MRI scans. The secondary objectives were to produce patient-specific three-dimensional reconstructions and to integrate these steps into a dedicated application, VenousNetworkAnalyzer (VNA).

## MATERIALS AND METHODS

The model was developed using a retrospective single-center cohort from a tertiary university hospital between 2019 and 2024. Among 225 MRI examinations initially screened, 31 were excluded due to insufficient image quality for reliable segmentation, resulting in a final cohort of 194 contrast-enhanced craniocervical MRI examinations (Figure 1). Exclusion criteria also included venous occlusive disease, cerebrospinal arteriovenous malformations or fistulas, prior posterior fossa or cervical spine surgery, and previous venotoxic treatments. Anatomical variations of the venous network were not considered exclusion criteria. The study was registered on ClinicalTrials.gov (NCT06815848). It was reported in accordance with the Checklist for Artificial Intelligence in Medical Imaging (CLAIM) 2024^7^.

**Figure 1.**
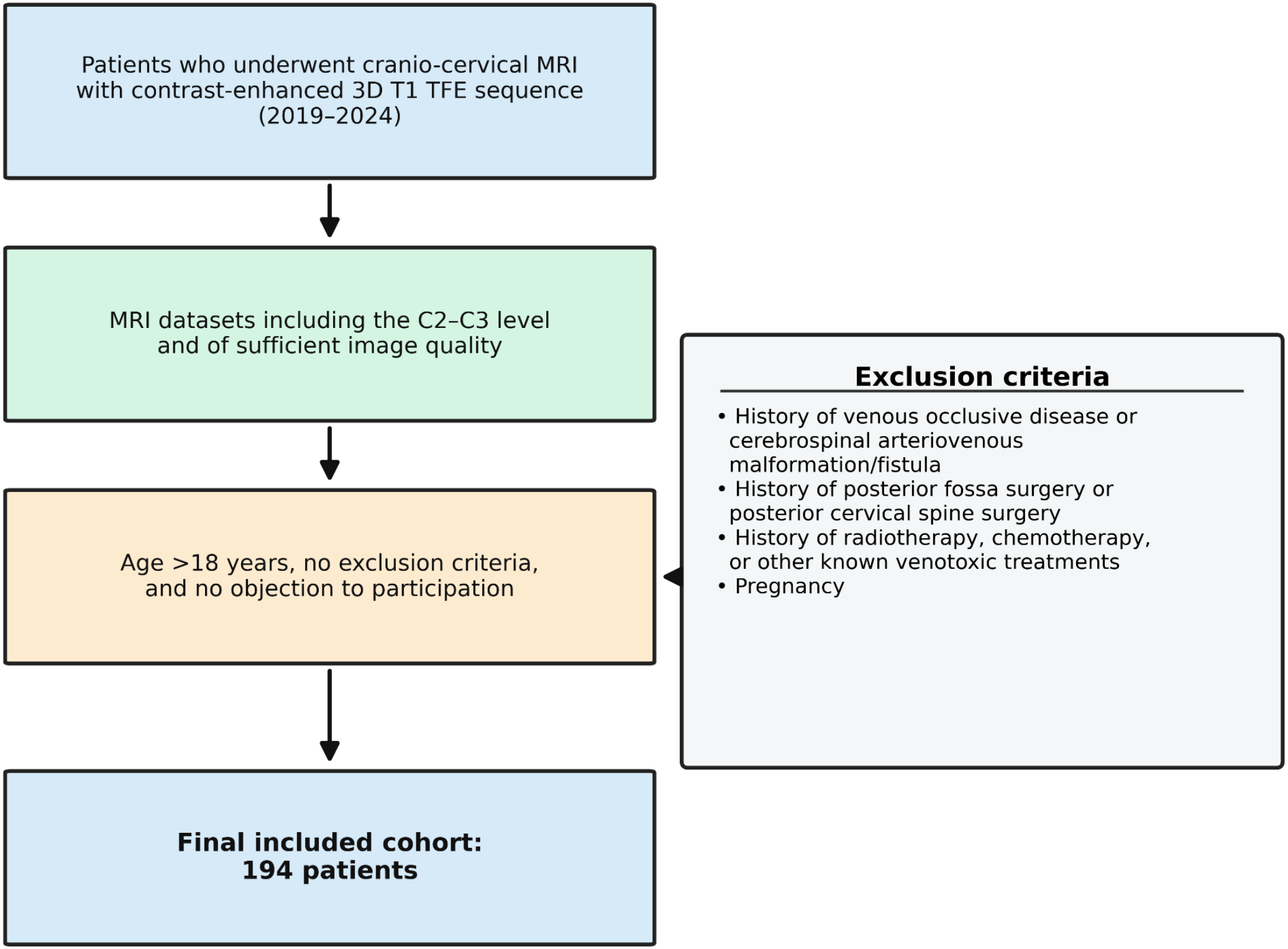
Flowchart of patient selection. This flowchart summarizes the selection process of patients included in the study cohort. Adult patients who underwent contrast-enhanced cranio-cervical MRI with a 3D T1 TFE sequence between 2019 and 2024 were screened. MRI datasets including the C2–C3 level were assessed for eligibility. Exclusion criteria are detailed on the right. The final included cohort consisted of 194 patients.

The scans were acquired on a 3-T Ingenia Elition X MRI scanner (Philips Healthcare). The acquisition was based on a 3D T1 TFE sequence following gadolinium injection, with the following parameters: repetition time (TR) of 8.24 ms, echo time (TE) of 3.80 ms, flip angle of 8°, slice thickness of 1 mm, acquisition matrix of 480, pixel size of 0.5 × 0.5 mm, and a volume comprising 340 slices. Imaging data were analyzed after anonymization

### 1. Development of the initial model

Automatic segmentation was developed using nnU-Net v2, an artificial intelligence tool specifically designed for medical image segmentation^8,9^. To create the initial dataset, manual reference segmentations were carried out in the 3D Slicer software (version 5.10.0) using the Segment Editor tool. All manual reference segmentations, including those of the vertebral artery, as well as all subsequent corrections of the automatic segmentations, were performed by a single author (a neurosurgery resident experienced in 3D Slicer). A structure was considered venous when it showed a tubular appearance, was visible on at least three consecutive slices, had an anatomically plausible location, and was continuous with an identifiable venous structure, such as a dural venous sinus or the internal jugular vein. Segmentation covered the entire MRI acquisition volume.

The order in which the scans were processed was randomized, and each scan was segmented only once to minimise any recall bias. Creating this initial dataset required approximately 10 hours of work per patient and involved 50 MRI scans. The models were trained over 500 epochs.Training and inference were performed on a workstation equipped with an Intel Core i7-14700 processor, 32 GB of RAM, and an NVIDIA H100 GPU with 80 GB of VRAM. Experiments were conducted under Linux using Python 3.11.9, PyTorch 2.10.0+cu128, and nnU-Net v2.6.4.

### 2. Optimisation of the segmentation model

An iterative strategy was adopted to gradually improve the model’s performance.Following each training step, the resulting model was applied to new MRI examinations; generated segmentations were systematically reviewed and manually corrected before being reintegrated into the training dataset for the subsequent model. Following this iterative process, all 194 segmentations were manually verified before subsequent analyses (Figure 2).

**Figure 2:**
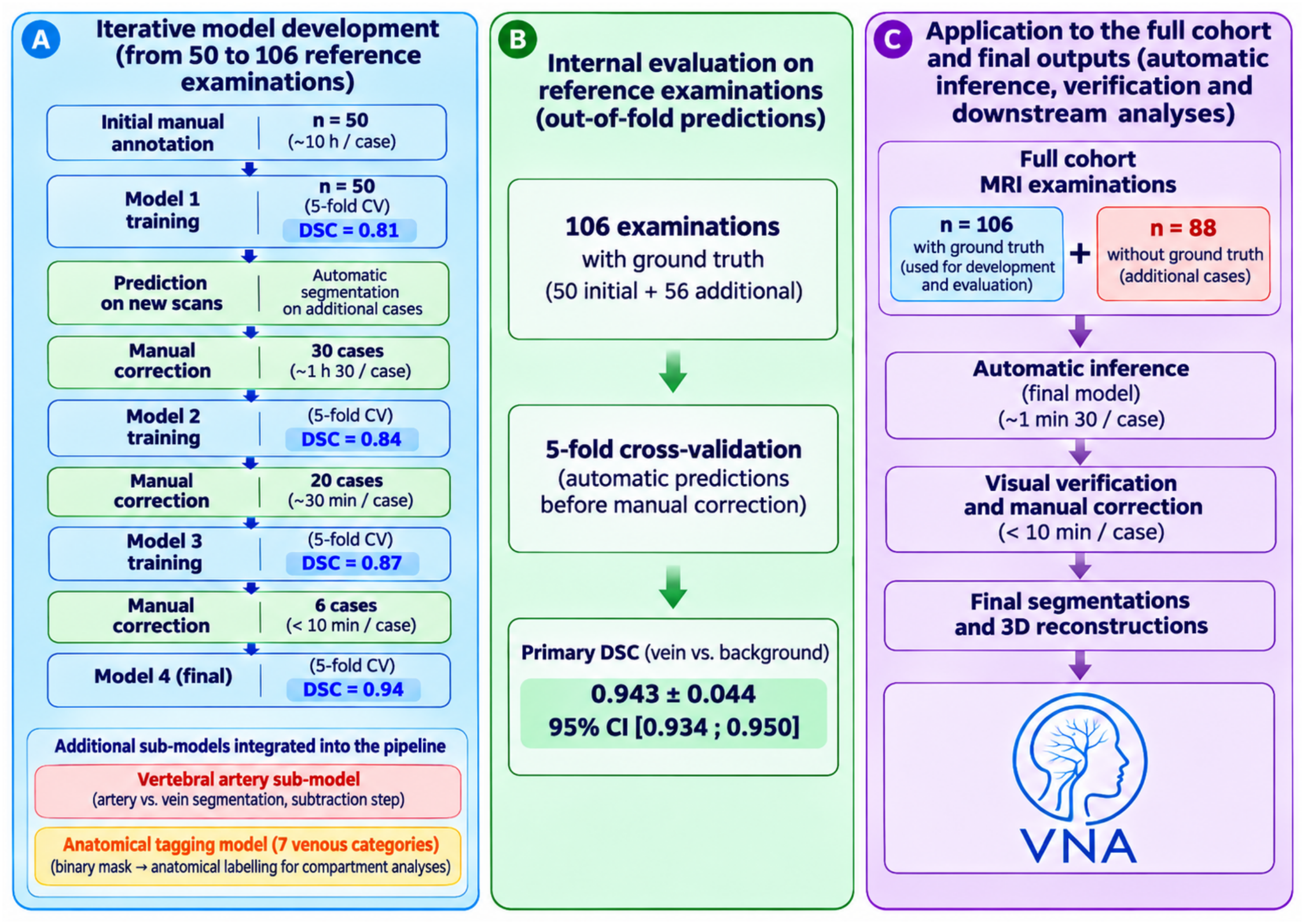
Iterative development workflow of the segmentation pipeline. Abbreviations: CCVS, cranio-cervical venous system; CI, confidence interval; CV, cross-validation; DSC, Dice similarity coefficient; GT, ground truth; MRI, magnetic resonance imaging; VNA, VenousNetworkAnalyzer.

Segmentation performance was assessed using the Dice similarity coefficient (DSC), which measures the spatial overlap between automatic and reference segmentations.In accordance with the recommendations of Bradshaw et al., segmentation performance was evaluated on 106 MRI examinations with available reference segmentations (50 fully manual and 56 obtained by manual correction of earlier model outputs) using five-fold internal cross-validation^10^. The dataset was divided into five groups, each successively used for evaluation while the remaining four were used for training. This approach aims to prevent direct overlap between training and evaluation data, thereby limiting the risk of overoptimistic DSC estimates due to overfitting and providing an internal estimate of model generalizability. DSC was calculated by comparing each uncorrected automatic prediction with the corresponding reference segmentation. To complement this volumetric overlap assessment, centerline Dice (clDice) was calculated to evaluate the agreement between the centerlines of automatic and reference segmentations. Three-dimensional skeletonization was performed on binary masks at native image resolution, and clDice was calculated as the harmonic mean of topology precision and topology sensitivity, as described by Shit et al^11^. Both metrics were calculated on the same 106 out-of-fold predictions generated without test-time augmentation.

Results were reported as mean ± standard deviation, with 95% confidence intervals calculated using bootstrap resampling with 10,000 iterations in Python. A separate descriptive analysis was performed by comparing predictions generated by the final segmentation pipeline with the manual reference segmentations of the same 106 examinations. Because these examinations had contributed to model development, this analysis was not considered an independent validation of segmentation performance. Additional volumetric agreement was assessed in the descriptive analysis using two indices adapted from radiosurgical conformity metrics. Letting VA denote the automatic segmentation volume, VR the manual reference volume, and VH their intersection, the coverage index (SI) was defined as SI = VH/VR, and the volumetric ratio (VRat) as VRat = VA/VR. The coverage index quantifies the proportion of the reference segmentation identified by the model, whereas the volumetric ratio compares total segmented volumes independently of their spatial overlap.Because the CCVS contains small, tortuous, and anatomically complex venous structures, quantitative evaluation was supplemented by systematic manual anatomical verification.

After model development and internal validation, the final model was applied to the remaining 88 examinations. These segmentations were visually inspected and minimally corrected when necessary before being used for three-dimensional reconstruction. Because no independent manually segmented reference was available for these 88 examinations, they were not included in the quantitative DSC analysis. Therefore, the reported performance should be interpreted as internal validation rather than external validation of model generalizability.

### 3. Exclusion of the vertebral artery

Segmenting the venous plexus of the vertebral artery presented an anticipated methodological challenge, due to its periarterial and sometimes plexiform organisation around the vertebral artery, particularly at the V3 segment. On contrast-enhanced 3D T1 sequences, the anatomical proximity and signal characteristics of these two structures—which are sometimes similar—posed a risk of arterial inclusion in the venous segmentation.

To minimise this risk, a specific step to exclude the vertebral artery was incorporated into the pipeline. Manual segmentation of the vertebral artery was performed on 50 scans where the arteriovenous distinction was unambiguous, requiring approximately two additional hours per patient. These data were used to train a sub-model designed to identify and then subtract the vertebral artery prior to the final segmentation of the venous network.

### 4. Anatomical labeling of the venous network

To facilitate anatomical interpretation of the three-dimensional reconstructions, the pipeline also includes an anatomical labeling step applied downstream of the binary CCVS segmentation. This step assigns each segmented venous voxel to one of seven predefined anatomical categories: intracranial venous structures (IC), internal jugular veins (IJV), external jugular veins (EJV), the vertebro-epidural compartment (VEC), including the vertebral artery venous plexus and epidural venous plexuses, deep cervical veins (DCV), the muscular venous network (M), and the subcutaneous venous network (SC). These categories were defined on anatomical grounds. The labeling model was trained with nnU-Net v2 on 30 manually labeled reconstructions; its development and compartment-specific evaluation are reported separately (manuscript in preparation). In the present study, anatomical labeling was used for visualization purposes only, and all quantitative results refer to the binary CCVS segmentation.

### 5. Integration into VenousNetworkAnalyzer

The entire pipeline has been integrated into a dedicated open-source application, VenousNetworkAnalyzer (VNA; version 1.0, developed by the authors). This application has been designed to bring together, within a single interface, the steps previously carried out separately: importing DICOM data, pre-processing, running the segmentation model, visual inspection of the resulting masks, and three-dimensional visualization of the reconstructed venous network. Input DICOM data were converted to NRRD format before segmentation (using SimpleITK). Output segmentations were exported as NRRD labelmaps for visual inspection in the application and in 3D Slicer, and for three-dimensional reconstruction using interactive in-browser rendering of the segmented voxels.

## RESULTS

### 1. Development of the segmentation pipeline

A total of 194 3D T1-weighted MRI scans with contrast were analyzed. The main demographic characteristics of the study sample are summarized in Figure 3. The development of the segmentation pipeline was based on an iterative process combining manual segmentation, model training, manual correction and retraining. The distribution of cases used during the various stages of development is shown in Figure 2.

**Figure 3:**
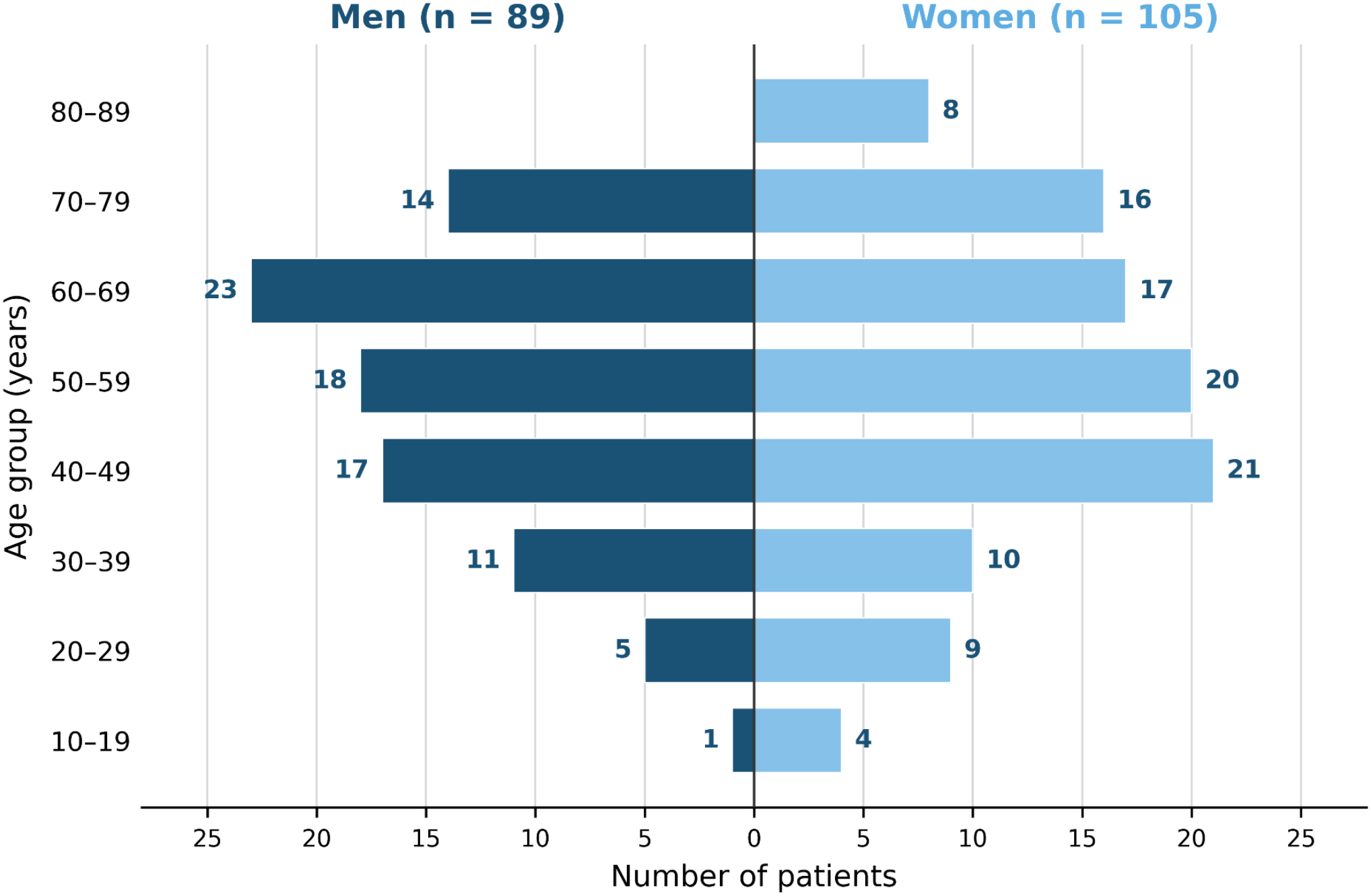
age and sexe distribution of the cohort (n=194)

Correction time progressively decreased from approximately half a day per patient with the first model to 1 hour 30 minutes with the second model, 30 minutes with the third model, and less than 10 minutes with the fourth model. Approximate training time was 72 hours for model 1, 65 hours for model 2, 50 hours for model 3, and 42 hours for model 4 (Supplementary Figure S1). With the final pipeline, automatic inference required approximately 1 min 30 s per examination. Subsequent visual verification and manual correction, when necessary, required approximately 8–8.5 minutes, resulting in a total processing time of approximately 10 minutes per examination, compared with approximately 10 hours for fully manual segmentation(Figure 2).

In five-fold internal cross-validation on the 106 examinations with available reference segmentations, the model achieved a mean DSC of 0.941 ± 0.044 (95% CI, 0.932–0.948) and a mean clDice of 0.917 ± 0.057 (95% CI, 0.906–0.927). The mean difference between clDice and DSC was −0.023. In a separate descriptive analysis of cohort predictions on the same 106 examinations, the mean DSC was 0.960 ± 0.026 (Figure 6A). As these examinations had contributed to model development, this second result was not considered an independent validation of segmentation performance. Additional volumetric analysis of the final-model predictions showed a mean coverage index (SI) of 0.958 ± 0.033 and a mean volumetric ratio (VRat) of 0.995 ± 0.026. Overall, 103 of 106 examinations (97%) showed an automatic-to-reference volume ratio within 5% of unity (Figure 6B).

### 2. Three-dimensional reconstructions of the craniocervical junction venous system

Applying the final model to the 194 scans enabled the production of individualised three-dimensional reconstructions of the venous system at the craniocervical junction for all patients in the cohort (Figure 4). These reconstructions enabled visual assessment of the segmented venous network and provided the basis for subsequent volumetric analyzes.

**Figure 4.**
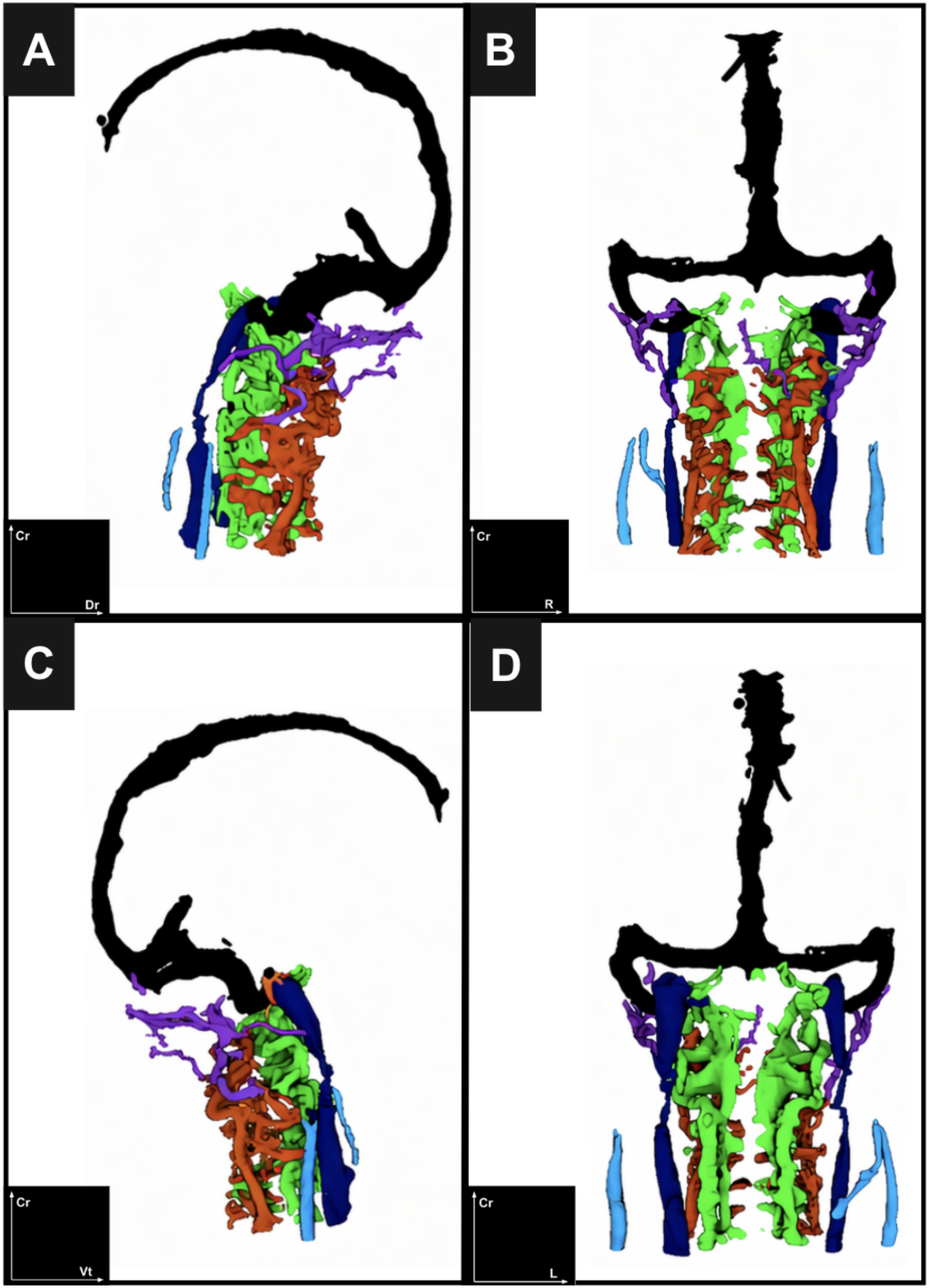
Three-dimensional reconstructions of the cranio-cervical venous system generated by the final segmentation model. (A) left lateral view, (B) posterior view, (C) right lateral view, and (D) anterior view. CR, cranial; DR, dorsal; R, right; VT, ventral; L, left.

**Figure 5.**
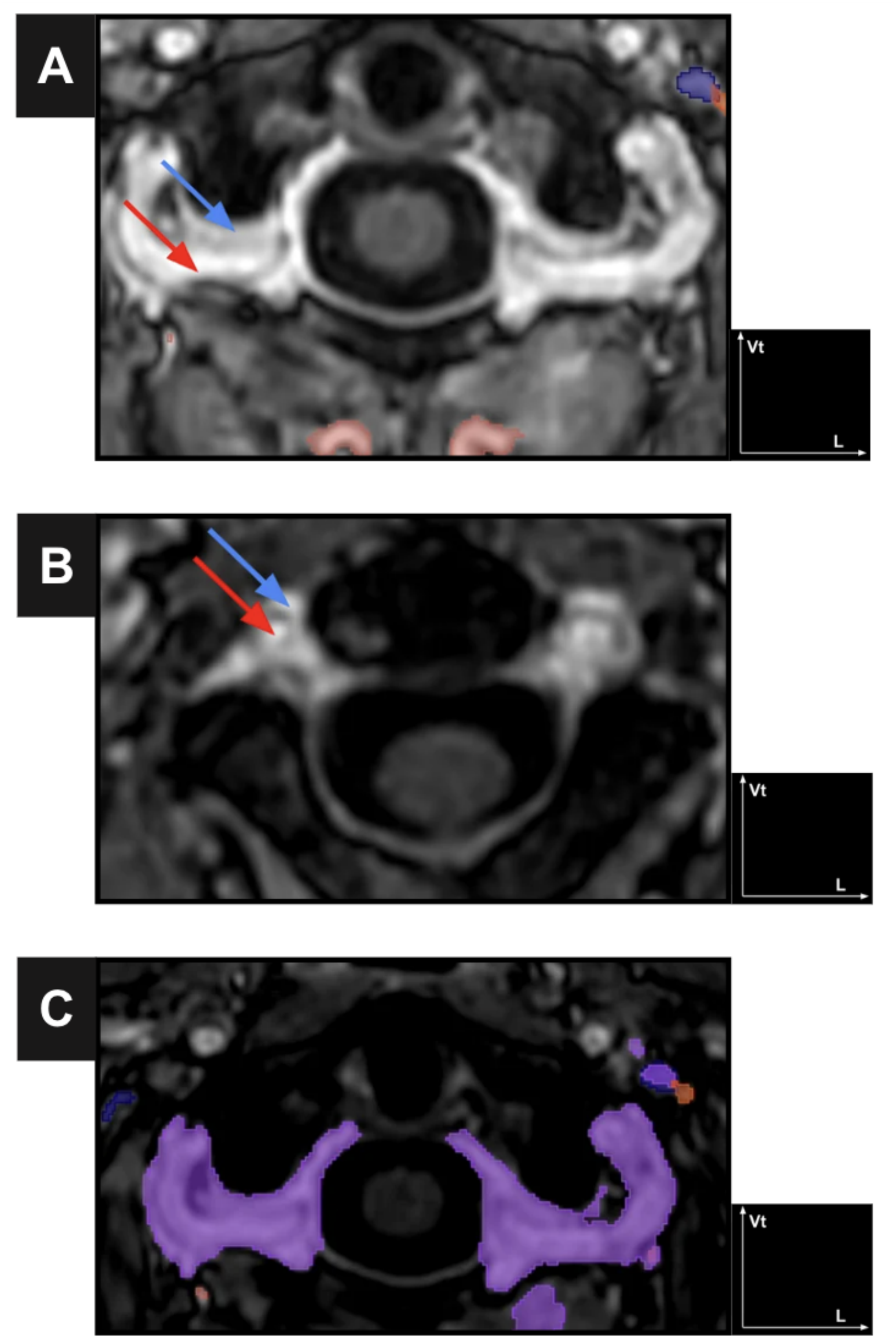
Segmentation difficulty involving the vertebral artery venous plexus. Axial gadolinium-enhanced T1 TFE MRI sections showing the close relationship between the vertebral artery venous plexus and the vertebral artery. Blue arrows indicate the venous plexus; red indicates the vertebral artery. Panel C shows arterial and venous mixing in an early segmentation model. **Abbreviations:** CR, cranial; VT, ventral; L, left.

**Figure 6.**
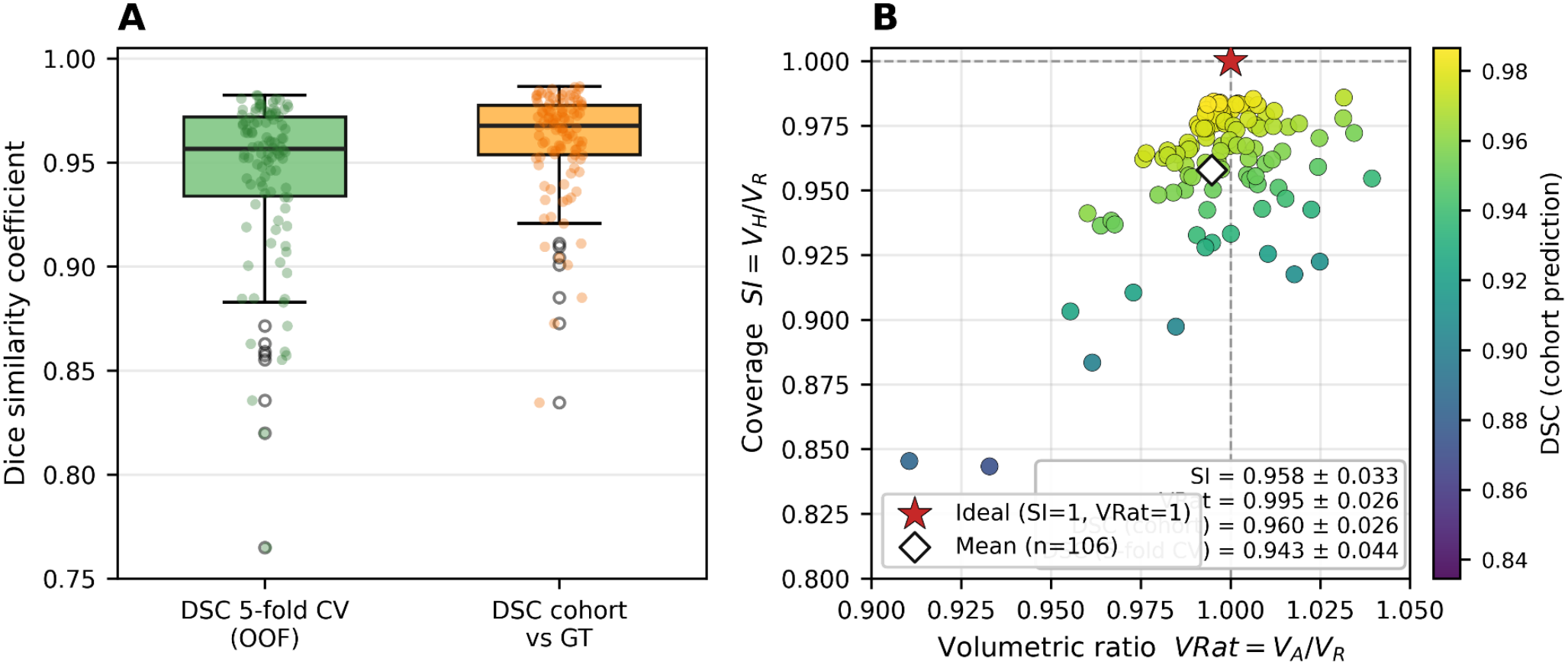
Segmentation performance and volumetric agreement. **(A)** Distribution of Dice similarity coefficients (DSC) across 106 examinations. Green indicates five-fold cross-validation predictions, and orange indicates cohort predictions. Points represent individual examinations, boxes the interquartile range, and horizontal lines the median. **(B)** Coverage index (SI = VH/VR) and volumetric ratio (VRat = VA/VR) for the same 106 examinations. Point color indicates cohort DSC. The red star represents ideal agreement, and the diamond represents the mean. **Abbreviations:** CV, cross-validation; DSC, Dice similarity coefficient; OOF, out-of-fold; SI, coverage index; VRat, volumetric ratio; VA, automatic volume; VR, reference volume; VH, intersection volume.

## DISCUSSION

### 1. Limitations of existing segmentation approaches

The development of a dedicated model was motivated by the limitations of currently available segmentation approaches for the CCVS. As summarized in Table 1, vascular enhancement filters, threshold-based segmentation, MIP reconstructions, and general-purpose segmentation tools may provide useful visual or anatomical information, but they are not designed to generate individualized, reproducible three-dimensional segmentations of the cranio-cervical venous networks on contrast-enhanced 3D T1 MRI^12,13^. These limitations justify the development of a sequence-specific deep-learning pipeline trained on manual reference segmentations and progressively optimized through iterative correction.

**Table 1.**
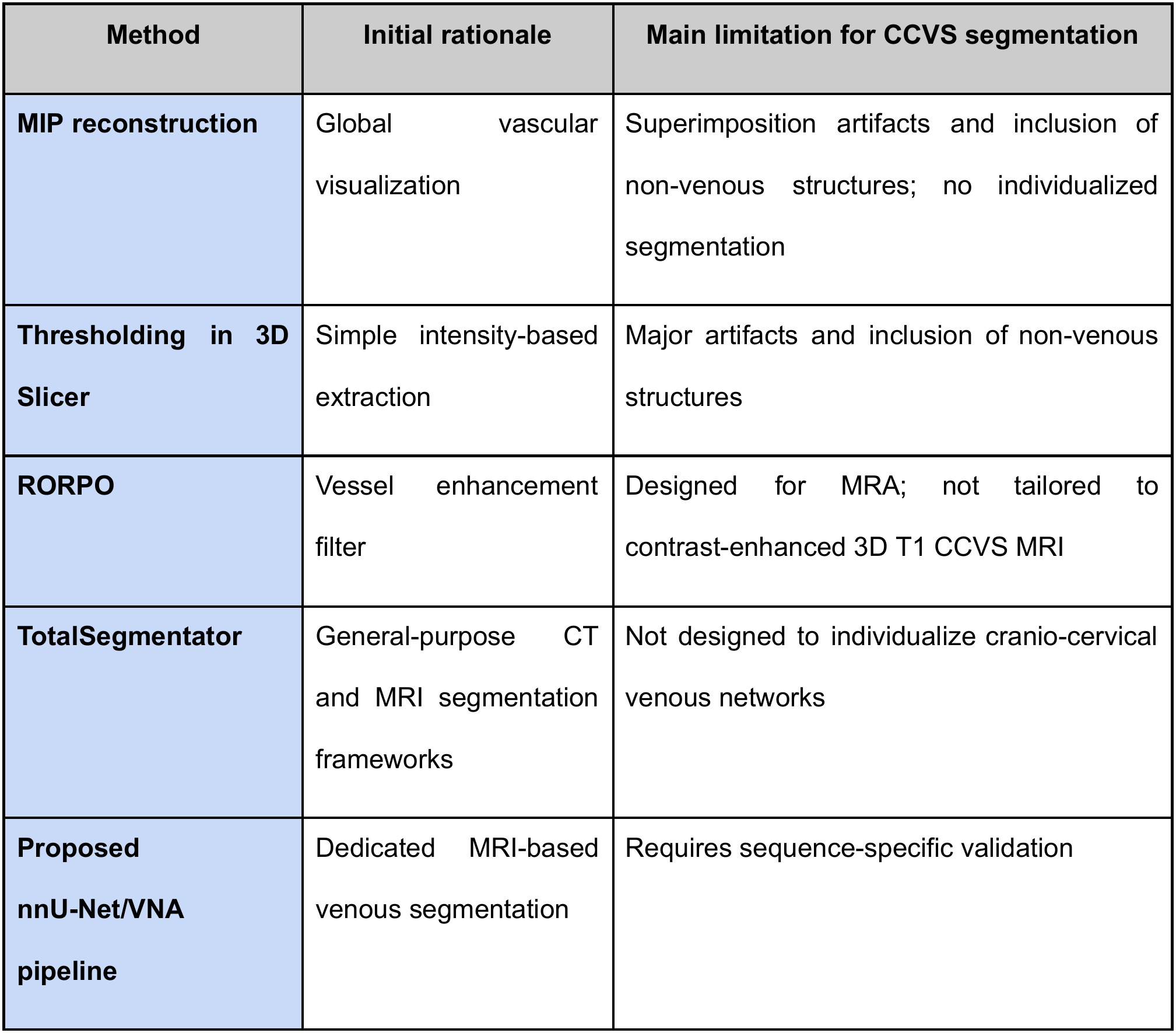
Comparison of available segmentation approaches for cranio-cervical venous system analyzis.

| **Method** | **Initial rationale** | **Main limitation for CCVS segmentation** |
|---|---|---|
| **MIP reconstruction** | Global vascular visualization | Superimposition artifacts and inclusion of non-venous structures; no individualized segmentation |
| **Thresholding in 3D Slicer** | Simple intensity-based extraction | Major artifacts and inclusion of non-venous structures |
| **RORPO** | Vessel enhancement filter | Designed for MRA; not tailored to contrast-enhanced 3D T1 CCVS MRI |
| **TotalSegmentator** | General-purpose CT and MRI segmentation frameworks | Not designed to individualize cranio-cervical venous networks |
| **Proposed nnU-Net/VNA pipeline** | Dedicated MRI-based venous segmentation | Requires sequence-specific validation |

### 2. Choice of imaging sequence

The choice of the gadolinium-enhanced T1-weighted TFE MRI sequence is justified by its characteristics, which are particularly well-suited to the study of the CCVS. It has therefore been evaluated in other applications, particularly in the context of cerebral venous thrombosis. This sequence offers several advantages, including rapid acquisition, excellent spatial resolution, increased sensitivity to slow-flowing blood and small vessels, and the ability to visualise venous structures regardless of haemodynamic conditions. It also allows for better definition of venous walls with sharper contours, combined with optimised signal-to-noise ratio and tissue contrast^14–16^.

### 3. Evaluation of segmentation performance

The DSC is a widely used metric for evaluating segmentation models in medical imaging^17,18^. Although the DSC obtained in this study indicates high volumetric agreement between automatic and reference segmentations, it does not directly assess vascular continuity. This limitation is particularly relevant to the CCVS, which contains fine, tortuous venous structures and small interconnecting vessels. The complementary clDice evaluation showed slightly lower agreement than conventional DSC, suggesting that some segmentation discrepancies may affect the centerlines of venous structures despite high overall volumetric overlap. However, clDice does not guarantee anatomically accurate vascular connectivity, and systematic manual anatomical verification remains necessary.

The DSC should not be interpreted as directly comparable to values reported by general-purpose segmentation frameworks such as TotalSegmentator, which was developed for CT-based segmentation and subsequently extended to MRI for multiple major anatomical structures^12,19^. By contrast, the present study focused on MRI-based segmentation of a fine venous network using reference segmentations performed by a single annotator. Surface-based metrics such as normalized surface Dice (NSD) could provide complementary information, but their interpretation depends on the selected tolerance threshold^12^.

### 3. Difficulties relating to the segmentation protocol

Two main difficulties were encountered during segmentation. The first concerned isolated venous segments, particularly within the muscular network, where early models produced discontinuous fragments requiring manual reconnection. The second concerned venous collapse variation near adjacent arterial structures, requiring manual identification of vascular continuity in early model versions.

### 4. Contribution of VNA

The integration of the workflow into VNA is an important step toward transferability. Unlike isolated scripts or technical environments that may be difficult to reproduce, VNA was designed to apply the model to new data, inspect the results, and generate three-dimensional reconstructions without advanced machine-learning expertise.

### 5. Limitations

Firstly, the data collected are exclusively morphological and non-dynamic; however, the presence of a vein and its caliber do not always determine its functional role in terms of blood flow and, consequently, its contribution to venous drainage^2,20,21^. Furthermore, there is a selection bias linked to the retrospective and hospital-based nature of the cohort, which does not include strictly healthy subjects. Additionally, all reference segmentations and corrections were performed by a single annotator without independent review, and inter-observer variability was not assessed. Moreover, 56 of the 106 reference segmentations were obtained by correcting model outputs rather than drawn from scratch, which may have slightly favored agreement with the automatic segmentations. Finally, another source of bias stems from the model’s dependence on the acquisition sequence, as it was developed and validated using a single contrast-enhanced 3D T1 TFE sequence. However, acquisition parameters frequently vary from one centre, or even one scanner, to another, and any modification could affect the quality of the automatic segmentation. The generalizability of the model to other acquisition conditions therefore remains to be assessed.

Despite these limitations, this study demonstrates the feasibility of automated MRI-based segmentation and patient-specific three-dimensional morphological reconstruction of the CCVS, supported by internal validation. The proposed pipeline substantially reduces processing time and provides a framework for further anatomical investigations. Future studies should assess its generalizability across different MRI acquisition protocols, investigate the functional contribution of individual venous pathways, and determine its potential clinical utility.

## Data Availability

The MRI datasets analyzed in this study are not publicly available due to patient confidentiality and institutional data protection requirements. Access to de-identified data may be considered upon reasonable request to the corresponding author, subject to institutional approval and applicable regulations. The VenousNetworkAnalyzer software and trained model weights are available through the project repository: https://github.com/marieduigou/VenousNetworkAnalyzer.

## Funding

This work was supported by a Master 2 research grant from the Journées de Neurologie de Langue Française.

## Competing interests

The authors declare that they have no relevant financial competing interests.

## Ethics approval

This retrospective study was conducted in accordance with the ethical standards of the institutional research committee and with the 1964 Declaration of Helsinki and its later amendments. Approval was obtained from the appropriate institutional ethics committee (study code: 29BRC24.0312).

## Consent to participate

In accordance with French regulations governing retrospective research involving previously acquired clinical imaging data, participants were informed of the use of their data for research purposes and were given the opportunity to object. No participant opposed the use of their data.

## Consent for publication

Not applicable. This manuscript does not contain identifiable individual participant data.

## Data sharing statement

Data generated or analyzed during the study are available from the corresponding author by request.

## Code and software availability

The VenousNetworkAnalyzer (VNA) software is publicly available at https://github.com/marieduigou/VenousNetworkAnalyzer. The version used in this study was VenousNetworkAnalyzer version 1.0.

## Author contributions

Marie Duigou contributed to study conception and design, data collection, manual segmentation, data analysis, interpretation of results, and manuscript drafting. Timothé Treguer contributed to software development, coding, and implementation of the machine learning pipeline. Brieg Dissaux, Julien Ognard, Romuald Seizeur, and Cyrille Capel contributed to study supervision, interpretation of results, and critical revision of the manuscript. All authors read and approved the final manuscript.

---

**Supplementary Figure S1.**
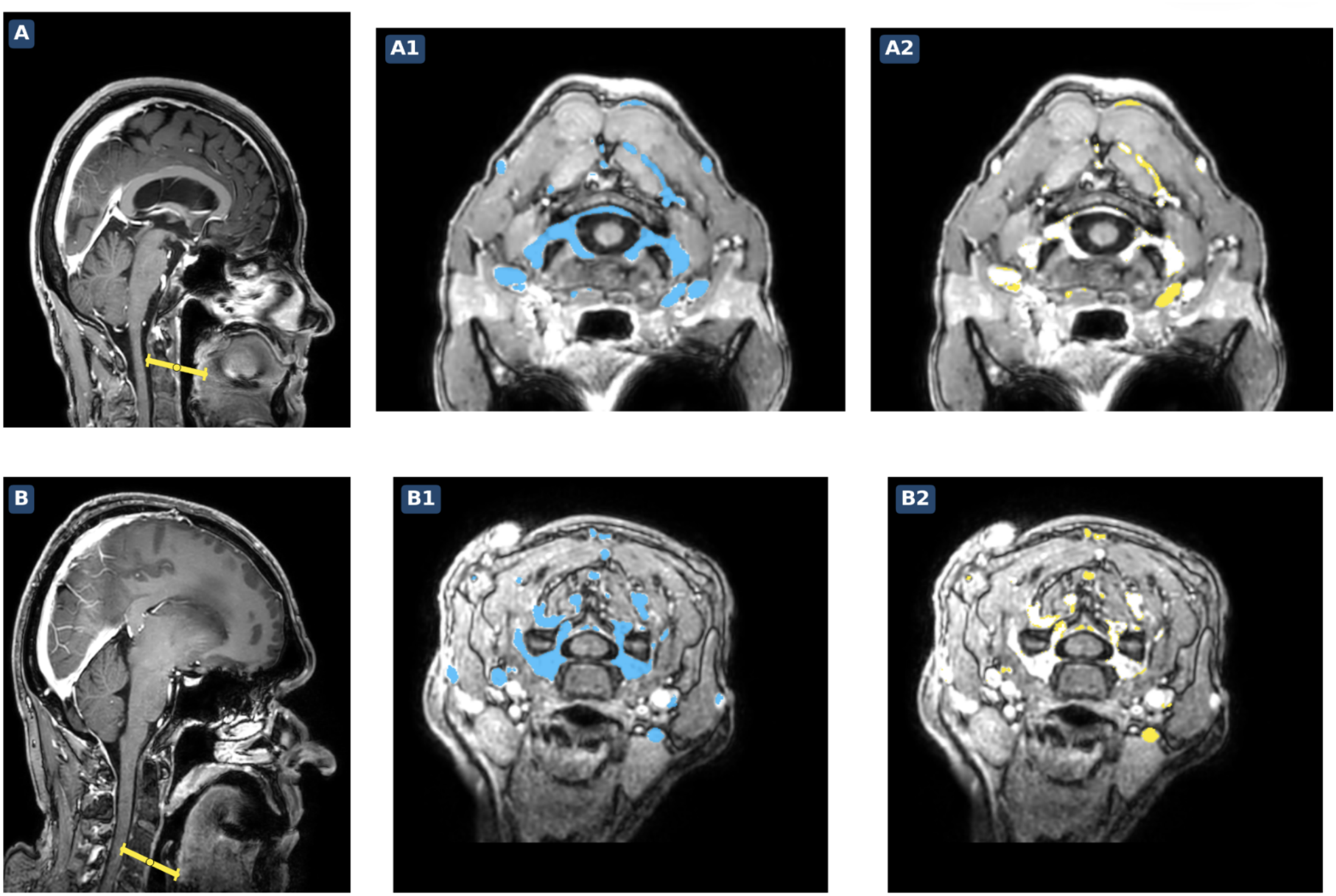
Representative false-negative segmentation errors during iterative model development. Two representative MRI examinations (A and B) illustrating segmentation errors identified after the second model, before manual correction. Sagittal localizers indicate the oblique axial imaging plane (yellow line). Corresponding axial images show the manual reference segmentation (A1, B1; cyan) and venous structures missed by the automatic model (A2, B2; yellow). These errors were subsequently corrected before training the third model.

## Notes

### Competing Interest Statement

The authors have declared no competing interest.

### Author Declarations

The study was reviewed by the Comite de Reflexion Ethique de Territoire of the Centre Hospitalier Universitaire de Brest (Brest University Hospital, France), under protocol reference 29BRC24.0312.

